# No impact of sex on surgical site infections in abdominal surgery: A multi-center study

**DOI:** 10.1101/2022.03.01.22271688

**Authors:** Simone Nora Zwicky, Severin Gloor, Franziska Tschan, Daniel Candinas, Nicolas Demartines, Markus Weber, Guido Beldi

## Abstract

**Objective:** Male sex is controversially discussed as a risk factor for surgical site infections (SSI). The aim of the present study was to evaluate the impact of sex on SSI in abdominal surgery under elimination of relevant confounders.

**Methods:** Clinicopathological data of 6603 patients undergoing abdominal surgery from a multi-center prospective database of four Swiss hospitals including patients between 2015 and 2018 were assessed. Patients were stratified according to postoperative SSI and risk factors for SSI were assessed using univariate and multivariate analysis.

**Results:** In 649 of 6603 patients SSI was reported (9.8%). SSI was significantly associated with reoperation (22.7% vs. 3.4%, p <0.001), higher mortality rate (4.6% vs. 0.9%, p <0.001) and higher rate of length of hospital stay over the 75^th^ percentile (57.0 % vs. 17.9 %, p <0.001). In univariate analysis male sex was a significant risk factor for SSI (p = 0.01). In multivariate analysis including multiple confounders ‘ such as comorbidities and perioperative factors there was no association between male sex and risk of SSI (odds ratio (OR) 1.1 [CI 0.8 – 1.4]). Independent risk factors for SSI in multivariate analysis were BMI ≥ 30 kg/m^2^ (OR 1.8 [CI 1.3 - 2.3]), duration of surgery > 75^th^ percentile (OR 2.3 [1.8 - 2.9]), high contamination level (OR 1.3 [1.0 – 1.6]), laparotomy (OR 1.3 [1.0 – 1.7]), pervious laparotomy (OR 1.4 [1.1 – 1.7]), blood transfusion (OR 1.7 [1.2 – 2.4]), cancer (OR 1.3 [1.0-1.8], malnutrition (OR 2.5 [1.8 – 3.4]).

**Conclusion:** Under elimination of relevant confounders there is no significant correlation between sex and risk of SSI after abdominal surgery.

## Introduction

Surgical site infections (SSI) rank among the three most frequent healthcare-associated infections in Europe. [1] They account for more than one fifth of all healthcare-associated infections with reported incidence of up to 10% and impose a substantial burden to both, health-care systems and patients. [1-6] In the past, multiple patient- and surgery-related factors contributing to the development of SSI were identified including age, body mass index (BMI) and duration of surgery. [6-10]

Recently, sex was intensively discussed as possible risk factor for SSI as several studies reported increased SSI rates for men. [11-15] For instance two high volume studies analysing data from German Nosocomial Infections Surveillance reported significantly higher incidence of SSI among men in different procedures including abdominal surgery. [11, 12] However, one study was not unadjusted for any confounders [11] and in the other important potential confounders as comorbidities including obesity and malnutrition could not be included into the analysis. [12]

Most other studies display unadjusted results or omit some important potential confounders as patient ‘s age, ASA score, obesity, malnutrition, grade of wound contamination, duration of surgery and surgical approach (e.g. laparotomy). [11-15] Therefore it remains unclear if male sex represents a risk factor for developing SSI after abdominal surgery.

The identification of sex as risk factor for SSI would be important to allow the generation of sex-specific infection control practices in the future. The aim of the present study is to evaluate the impact of sex on surgical site infections after abdominal surgery under elimination of possible confounders.

## Methods

### Trial design, patients, study interventions and data collection

The study was designed as a multi-center, retrospective analysis of an existing prospective collected database. This database was created in the context of a prospective trial that assessed the effects of structured intraoperative briefings on patient outcomes including SSI. [16] The database includes data from patients undergoing surgery in four Swiss hospitals (2 university hospitals, and two non-university referral centres) between May 2015 and March 2018. The database was initiated after obtaining approval from the respective local ethical committees (leading committee: Kantonale Ethikkommission Bern #161/2014). Written general consent was obtained from individuals of three centers, in the fourth center the local ethical committee allowed explicitly the inclusion of patients who did not refuse the use of their data. All patients with an indication for elective or emergency abdominal surgery in the participating hospitals were included and entered into an electronic database. Clinicopathological data of 6603 patients including demographics, therapeutic features, surgical procedures and complications as SSI were extracted. For the final analysis 82 patients with death within 30 days after surgery were excluded. All surgical treatments were performed by board-certified surgeons. Patients received in-hospital disease surveillance and therapy according to institutional standards and international guidelines. Exclusion criteria were patient age <18 years, preexisting SSI, previous surgery at the same surgical site within the past 30 days, procedures without general anaesthesia and proctological operations. Further patients with American Society of Anesthesiologists (ASA) - score ≥5 were excluded. [17]

### Study endpoints

The primary endpoint of this study was defined as SSI within 30 days after surgery according to the Center for Disease Control and Prevention criteria. [18] Trained study nurses evaluated SSIs in accordance with the Swissnoso SSI surveillance system guide. [19] This guide adheres to the US National Healthcare Safety Network (former National Nosocomial Infection Surveillance, NNIS) standards and includes follow-up interviews by telephone 30 days after surgery. [18, 20] One center entered their perioperative data to the Enhanced Recovery After Surgery Interactive Audit System (ERAS; Encare, Stockholm, Sweden) and SSI were evaluated based on this validated data set. [21]

### Statistical analysis

Quantitative and qualitative variables were expressed as mean (standard deviation) and frequency (percentage).For all outcomes, the two groups were compared with the Fisher exact test or Kruksal-Wallis test for categorical variables and the Mann-Whitney U test for continuous variables, as appropriate. All *p*-values are considered statistically significant if *p* < 0.05. Univariate analysis was applied for patient- and surgery-specific covariates to identify statistically significant association with the occurrence of SSI. Co-variates with statistically significant differences were considered as possible confounders for development of SSI. All confounders were eliminated with backward selection (*p*-value < 0.05). Statistical analysis was performed using SPSS version® 25 (IBM, Armonk, New York, USA).

## Results

A total of 6603 patients undergoing abdominal surgery between May 2015 and March 2018 were analysed. Mean age and BMI were significantly increased in the SSI group (62.0 vs 56.6 years, *p <0*.*001*; 27.5 vs 26.9 kg/m2, *p = 0*.*010*). Patient demographics and surgical characteristics are displayed in Table 1. Of the 6603 surgical interventions, 24.8% were colorectal operations (including small-bowel surgeries), 21.4% hernia repairs, 16% cholecystectomies, 11.0% hepato-pancreato-biliary surgery, 10.% appendectomies, 8.2% bariatric interventions, 5.6% upper-GI operations and 3.0% renal and adrenal surgery. Mean duration of surgery (203.9 vs. 121.5 min, *p <0*.*001*), National Nosocomial Infections Surveillance (NNIS) index (1.24 vs. 0.7, *p <0*.*001*) and ASA (2.6 vs. 2.2, *p <0*.*001*) score were significantly higher in the SSI group. [22] SSI correlated significantly with increased rates of long LOS >75^th^ percentile (57.0 vs 17.9%, *p <0*.*001*), reoperations (22.7 vs 3.4%, *p <0*.*001*) and mortality (4.6 vs 0.9%, *p <0*.*001*). In 649 of 6521 included patients SSI was identified (9.8%). Univariate and multivariate analysis of patient and procedure dependent risk factors for SSI are shown in Table 2.

**Table 1.** Patient demographic and surgical characteristics.

| Variable | SSI<br>(n=649) | No SSI<br>(n=5954) | *p-value* |
| --- | --- | --- | --- |
| Age, years, mean years (SD) | 62.0 (14.9) | 56.6 (17.5) | <0.001 |
| BMI, mean kg/m<sup>2</sup> (SD) | 27.5 (6.9) | 26.9 (6.5) | 0.011 |
| Male sex, n (%) | 399 (61.5) | 3367 (56.6) | 0.016 |
| ASA Score, mean (SD) | 2.6 (0.7) | 2.2 (0.7) | <0.001 |
| NNIS index, mean (SD) | 1.24 (0.88) | 0.77 (0.80) | <0.001 |
| Duration of surgery, mean<br>minutes (SD) | 203.97 (114.2) | 121.55 (88.7) | <0.001 |
| Type of surgery, n (%) |  |  | 0.002* |
| Colorectal | 292 (45.0) | 1346 (22.6) |  |
| Hepato-Pancreato-Biliary | 137 (21.1) | 592 (9.9) |  |
| Renal and Adrenal | 6 (0.9) | 192 (3.2) |  |
| Appendectomy | 23 (3.5) | 638 (10.7) |  |
| Cholecystectomy | 48 (7.4) | 1009 (16.9) |  |
| Hernia | 43 (6.6) | 1368 (23.0) |  |
| Bariatric | 39 (6.0) | 503 (8.4) |  |
| Upper GI | 61 (9.4) | 306 (5.1) |  |
| LOS (>75%), n (%) | 369 (57.0) | 1063 (17.9) | <0.001 |
| Reoperation, n (%) | 131 (22.7) | 185 (3.4) | <0.001 |
| 30-Day mortality, n (%) | 30 (4.6) | 52 (0.9) | <0.001 |

ASA, American Society of Anesthesiology; BMI, body mass index; LOS, length of stay; NNIS, National Nosocomial Infections Surveillance; SD, standard deviation; SSI, surgical site infection. \* Kruksal-Wallis test

**Table 2.**
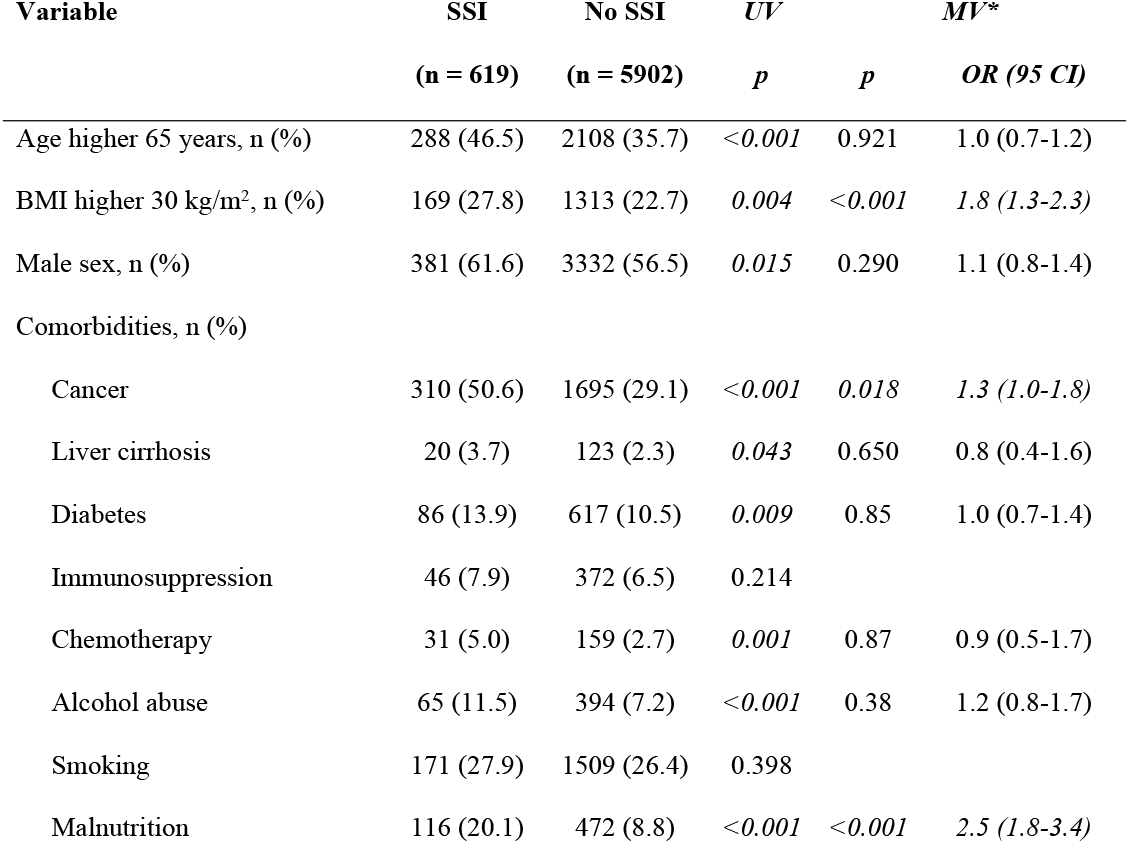
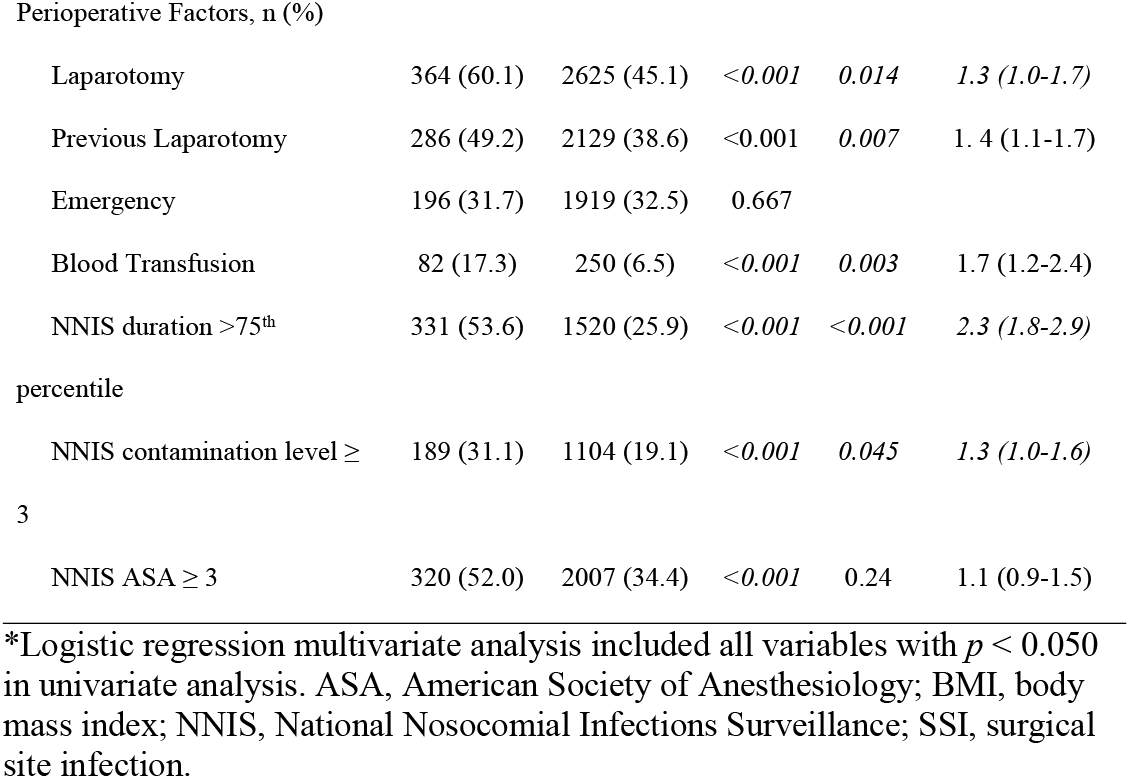
Univariate and multivariate analysis of patient and procedure dependent risk factors for SSI.

| Variable | SSI<br>(n = 619) | No SSI<br>(n = 5902) | UV<br>*p* | *p* | MV*<br>*OR (95 CI)* |
| --- | --- | --- | --- | --- | --- |
| Age higher 65 years, n (%) | 288 (46.5) | 2108 (35.7) | <0.001 | 0.921 | 1.0 (0.7-1.2) |
| BMI higher 30 kg/m<sup>2</sup>, n (%) | 169 (27.8) | 1313 (22.7) | 0.004 | <0.001 | 1.8 (1.3-2.3) |
| Male sex, n (%) | 381 (61.6) | 3332 (56.5) | 0.015 | 0.290 | 1.1 (0.8-1.4) |
| Comorbidities, n (%) |  |  |  |  |  |
| Cancer | 310 (50.6) | 1695 (29.1) | <0.001 | 0.018 | 1.3 (1.0-1.8) |
| Liver cirrhosis | 20 (3.7) | 123 (2.3) | 0.043 | 0.650 | 0.8 (0.4-1.6) |
| Diabetes | 86 (13.9) | 617 (10.5) | 0.009 | 0.85 | 1.0 (0.7-1.4) |
| Immunosuppression | 46 (7.9) | 372 (6.5) | 0.214 |  |  |
| Chemotherapy | 31 (5.0) | 159 (2.7) | 0.001 | 0.87 | 0.9 (0.5-1.7) |
| Alcohol abuse | 65 (11.5) | 394 (7.2) | <0.001 | 0.38 | 1.2 (0.8-1.7) |
| Smoking | 171 (27.9) | 1509 (26.4) | 0.398 |  |  |
| Malnutrition | 116 (20.1) | 472 (8.8) | <0.001 | <0.001 | 2.5 (1.8-3.4) |
| Perioperative Factors, n (%) |  |  |  |  |  |
| Laparotomy | 364 (60.1) | 2625 (45.1) | <0.001 | 0.014 | 1.3 (1.0-1.7) |
| Previous Laparotomy | 286 (49.2) | 2129 (38.6) | <0.001 | 0.007 | 1.4 (1.1-1.7) |
| Emergency | 196 (31.7) | 1919 (32.5) | 0.667 |  |  |
| Blood Transfusion | 82 (17.3) | 250 (6.5) | <0.001 | 0.003 | 1.7 (1.2-2.4) |
| NNIS duration >75<sup>th</sup> percentile | 331 (53.6) | 1520 (25.9) | <0.001 | <0.001 | 2.3 (1.8-2.9) |
| NNIS contamination level ≥ 3 | 189 (31.1) | 1104 (19.1) | <0.001 | 0.045 | 1.3 (1.0-1.6) |
| NNIS ASA ≥ 3 | 320 (52.0) | 2007 (34.4) | <0.001 | 0.24 | 1.1 (0.9-1.5) |

\*Logistic regression multivariate analysis included all variables with $p < 0.050$ in univariate analysis. ASA, American Society of Anesthesiology; BMI, body mass index; NNIS, National Nosocomial Infections Surveillance; SSI, surgical site infection.

In univariate analysis male sex was a significant risk factor for SSI (*p = 0*.*010*). Further patient dependent risk factors were age higher 65 years (*p <0*.*001*), BMI higher than 30 kg/m^2^ (*p = 0*.*004*), cancer (*p <0*.*001*), diabetes (*p = 0*.*009*), chemotherapy (*p = 0*.*001*), alcohol abuse (*p <0*.*001*) and malnutrition (*p<0*.*001*). Perioperative factors with significant correlation to SSI were open approach by laparotomy (*p <0*.*001*), previous laparotomy (*p <0*.*001*), type of surgical procedure (*p = 0*.*002*), blood transfusion (*p <0*.*001*) as well as all subsets of the NNIS score (duration>75^th^ percentile, contamination level ≥3 ASA grade ≥3, *p <0*.*001*). In multivariate analysis no association between male sex and risk of SSI (OR 1.1 [CI 0.8 – 1.4]) was seen. Figure 1 displays multivariate analysis, where BMI ≥ 30 kg/m^2^ (OR 1.8 [CI 1.3 -2.3]), cancer (OR 1.3 [1.0-1.8]), malnutrition (OR 2.5 [1.8 – 3.4]), duration of surgery > 75^th^ percentile (OR 2.3 [1.8 - 2.9]), contamination level ≥3 (OR 1.3 [1.0 – 1.6]), laparotomy (OR 1.3 [1.0 – 1.7]), pervious laparotomy (OR 1.4 [1.1 – 1.7]) and blood transfusion (OR 1.7 [1.2 – 2.4]) were persistent independent risk factors for SSI.

**Figure 1.**
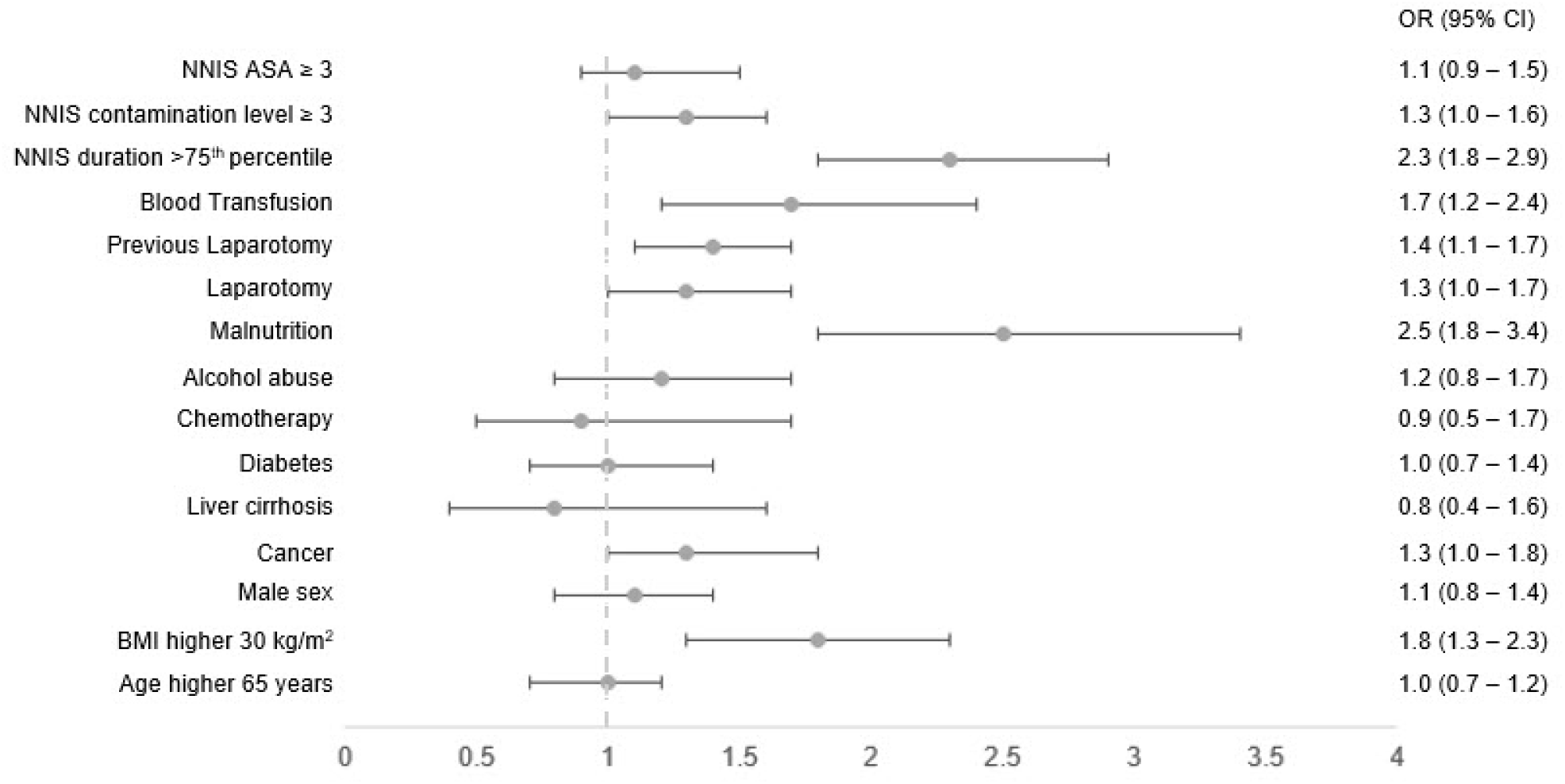
Multivariate analysis of factors associated with SSI following abdominal surgery in the entire study cohort.

## Discussion

Male sex is controversially discussed as a risk factor for SSI after abdominal surgery. Our study demonstrates no significant correlation between male sex and SSI in abdominal surgery under elimination of multiple relevant confounders. Age older than 65 years, a BMI over 30 kg/m^2^, an ASA score over 2 as well as a NNIS index over 2 correlated significantly with risk of SSI.

In contrast to our study, several high volume studies postulate higher risk of SSI for male compared to female patients after abdominal surgery. [11-15] However, the data were either presented completely unadjusted for potential confounders or some important confounders and comorbidities, as duration of surgery, obesity, malnutrition, were not be included into the analysis. [11-15] In particular, two studies demonstrating higher risk for SSI for men either displaying unadjusted results [11] or potential confounding through comorbidities as obesity, malnutrition as well as wound contamination could not be excluded. [12] Similarly male sex was identified as independent risk factor for SSI after gastric and pancreatic surgery, however factors as obesity and malnutrition [14] or wound contamination level [13] were omitted in multivariate analysis. A study addressing the risk of SSI after cholecystectomy, reporting significantly increased risk of SSI for male patients, omitted potential important risk factors as duration of surgery or wound contamination level in the multivariate analysis.[15] The study further demonstrated increased rate of severe biliary disease for men. A possible residual confounding caused by incomplete capture of the increased rate of severe biliary disease in men using claims data is discussed in the study. [13, 15]

The studies discussed different potential causes for the higher rate of SSI in men. [11-13] Higher rate of visceral fat in men may cause higher complexity of surgery and offer optimal conditions for bacterial growth. [12, 13, 23] Further hormonal or evolutional causes were postulated. [11, 12] However, in all of the named studies at least one potential important risk factor for SSI as malnutrition, obesity, duration of surgery or wound contamination level was omitted. [11-15] The potential impact of the missing patient as well as surgery-related confounders on the postulated higher risk of SSI in these studies therefore remains unknown. [11-15]

In the present study we demonstrate obesity, malnutrition, cancer, perioperative blood transfusion, long duration of surgery, high contamination level, laparotomy and previous surgery as independent significant risk factors for SSI. All of the named factors are widely-established and show the comparability of our treated patients to other centers. The association of obesity and surgical site infection was shown in several studies, the causality seems multifactorial. [24] It is well-known that malnutrition comes along with immune dysfunction, impaired wound-healing and SSI. [25-28] Increased risk of SSI for patients with cancer may likely be explained due to immunosuppressive state caused by the oncological disease as well as its treatment. [29, 30] The association of long duration of surgery > 75^th^ percentile with SSI is consistent with the finding of multiple studies. The effect may be caused by different factors including association with more complex surgery and increased fatigue of the surgical team leading to more technical mistakes. [7, 30, 31] Higher risk of SSI for patients with contaminated or dirty wound levels is commonly known. [32] The significant correlation of blood transfusion with risk of SSI is consistent to several studies. [33-36] On one hand need of blood transfusion indicates complex or maybe even uncontrolled intraoperative situation on the other hand immunosuppressive effects of blood transfusions are known. [34, 37] Higher rates of SSI for patients with open surgical approach as well as previous laparotomy are likely explained due to association with increased surgical difficulty, more extended surgeries and higher risk of organ lesion. [31, 38-41]

Despite the propective collection of the data, this study is limited by the retrospective design and therefore was not able to collect all potential sex-associated factors. Therefore, an effect caused by absence of potiental further confounders cannot be excluded. Because of the same reasons, aspects of gender were not included in this study. However, the most relevant strength of the present study is the assessment of a wide variety of potential risk factors, including all of the most commonly established risk factors for development of SSI allowing even multivariate analysis.

## Conclusion

After adjustment for wide variety of potential confounders including patient-related and procedure-related factors we could not correlate sex with rate of SSI. However, BMI≥ 30 kg/m^2^, duration of surgery > 75^th^ percentile, high contamination level, surgical approach, cancer, blood transfusion and malnutrition are significant independent risk factors for SSI in abdominal surgery and should be taken into account to reduce rate of SSI in abdominal surgery in the future.

## Data Availability

All relevant data are within the manuscript and its Supporting Information files. Original data (full data set) is made available upon request.

## Acknowledgements

None.

